# Mild Behavioral Impairment and Subjective Cognitive Decline predict Mild Cognitive Impairment

**DOI:** 10.1101/2020.05.24.20112284

**Authors:** Zahinoor Ismail, Alexander McGirr, Sascha Gill, Sophie Hu, Nils D. Forkert, Eric E. Smith

## Abstract

**Objective:** Better methods for detecting preclinical neuropathological change are required for prevention of dementia. Mild behavioral impairment (MBI) and subjective cognitive decline (SCD) can represent neurobehavioral and neurocognitive axes of early stage neurodegenerative processes, which are represented in Stage 2 of the NIA-AA Alzheimer’s disease research framework. Both MBI and SCD may offer an opportunity for premorbid detection. We test the hypothesis that MBI and SCD confer additive risk for incident cognitive decline.

**Methods:** Participants were cognitively normal older adults followed up approximately annually at Alzheimer’s Disease Centers. Logistic regression was used to determine the relationship between baseline classification (MBI+, SCD+, neither (MBI-SCD-), or both (MBI+SCD+)) and cognitive decline, defined by Clinical Dementia Rating (CDR) total score, at 3 years.

**Results:** Of 2769 participants (mean age=76; 63% females), 1536 were MBI-SCD-, 254 MBI-SCD+, 743 MBI+SCD-, and 236 MBI+SCD+. At 3-years, 349 individuals (12.6%) developed cognitive decline to CDR>0. Compared to SCD-MBI-, we observed an ordinal progression in risk, with ORs [95% CI] as follows: 3.61 [2.42-5.38] for MBI-SCD+ (16.5% progression), 4.76 [3.57-6.34] for MBI+SCD-, (20.7% progression) and 8.15 [5.71-11.64] for MBI+SCD+ (30.9% progression).

**Conclusion:** MBI in older adults alone or in combination with SCD is associated with a higher risk of incident cognitive decline at 3 years. The highest rate of progression to MCI is observed in those with both MBI and SCD. Used in conjunction, MBI and SCD could be simple and scalable methods to identify patients at high risk for cognitive decline for prevention studies.

## Introduction

Commonly cited reasons for high costs and poor outcomes in Alzheimer’s disease (AD) clinical trials are screen failures and poor recruitment of early phase illness^1,2^. Identification of sensitive and specific premorbid indicators of emergent pathology is a priority^1^. A leading strategy to detect preclinical disease is to focus on subjective cognitive decline (SCD), a perceived decline in cognitive ability in the absence of objective findings^3,4^, which has been associated with amyloid burden^5^ and incident cognitive decline and dementia^6^.

An emerging strategy is to capture early behavioral manifestations of dementia^7^ that occur in 30% of AD patients prior to cognitive manifestations^8^. Mild behavioral impairment (MBI) is a validated syndrome that serves as a sensitive transitional state marker for dementia syndromes. MBI is characterized by the emergence in later life of persistent neuropsychiatric symptoms, and may be an index manifestation of dementia, evident before cognitive symptoms^9^. MBI is associated with cognitive decline^10^, faster progression to dementia^11,12^, known AD genetic loci^13,14^, greater axonal loss^15^, and amyloid burden in patients with normal cognition^16^.

Both MBI and SCD are reflected in the NIA-AA AD research framework in Stage 2 (**Table 1, Figure 1**)^17^. To our knowledge, there have been no large prospective studies examining the prognostic utility of MBI and SCD in a sample of objectively normal individuals at higher risk for dementia. We hypothesized that cognitive and behavioral changes in late life may represent coherent or divergent manifestations of emerging pathology that can be leveraged to identify sensitive windows for intervention.

**Table 1:** Representation of SCD and MBI in NIA-AA Research Framework Clinical Stage 2^17^

| NIAA-AA Stage 2 Descriptors | SCD or MBI criteria |
| --- | --- |
| Transitional cognitive decline: Cognitive performance in the nonimpaired range but with a subjective complaint of cognitive decline, or a subtle decline measured on longitudinal cognitive testing, or neurobehavioral symptoms, or combinations of these | **SCD:** Self-experienced persistent decline in cognitive capacity in comparison with a previously normal status and unrelated to an acute event, normal age-, gender-, and education-adjusted performance on standardized cognitive tests<br>**MBI:** Behavior and personality changes can precede cognitive decline and present in absence of cognitive changes, or can accompany cognitive symptoms |
| Represents a change from individual baseline within past 1–3 years, and persistent for at least 6 months. Although cognition is the core feature, mild neurobehavioral changes—for example, changes in mood, anxiety, or motivation—may coexist. In some individuals, the primary complaint may be neurobehavioral rather than cognitive. Neurobehavioral symptoms should have a clearly defined recent onset, which persists and cannot be explained by life events | **SCD:** Onset of cognitive symptoms within the last 5 years (SCD-plus criterion<sup>3</sup>)<br>**MBI:** Changes in behavior or personality, starting later in life representing a clear change from usual behavior or personality, and persisting for at least 6 months; not better accounted for by psychiatric conditions (including adjustment difficulties secondary to life events) |

**Figure 1:**
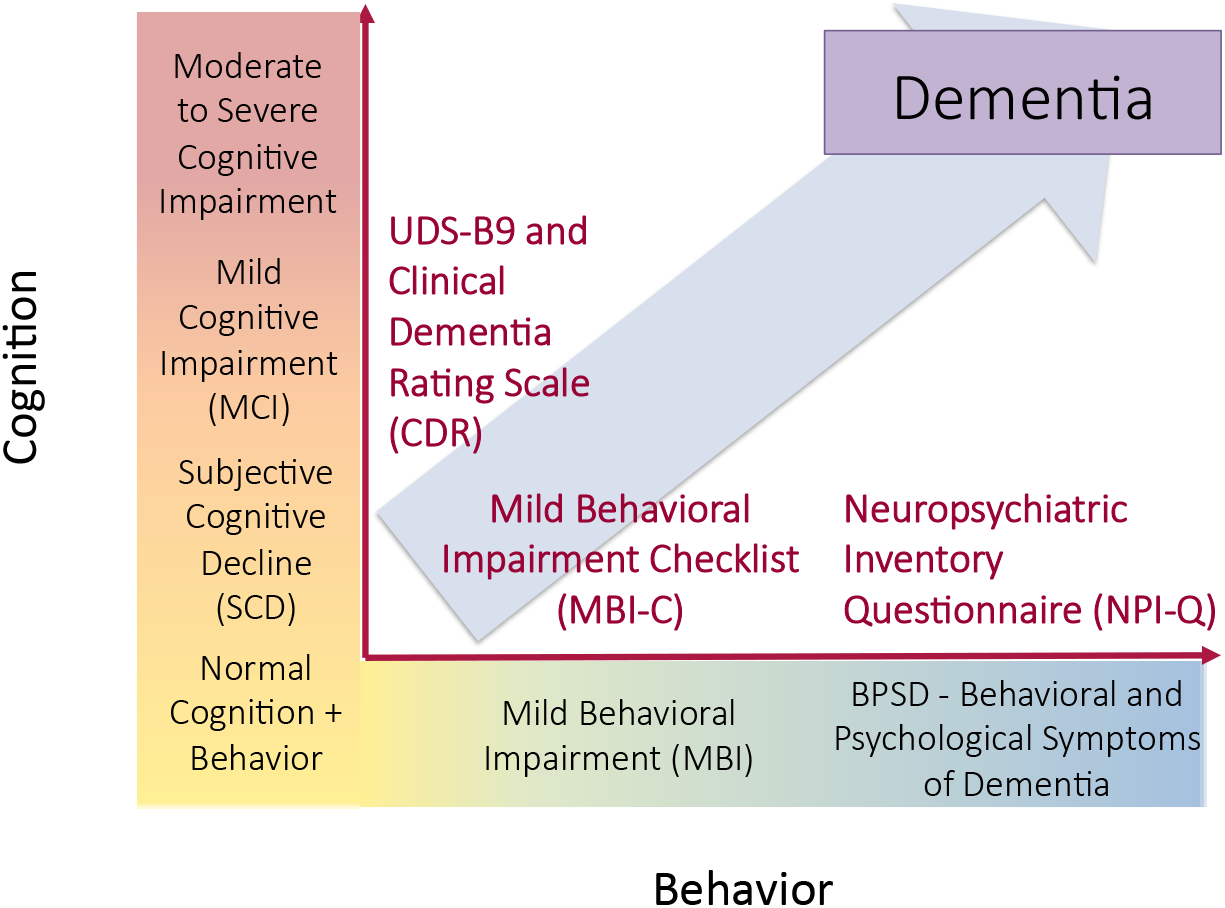
Cognitive and behavioral pre-dementia risk axes

## Methods

### Source population: National Alzheimer’s Coordinating Center (NACC)

Data used in this study were obtained from the NACC database (https://www.alz.washington.edu/). NACC was established by the National Institute on Aging (NIA) and consists of multiple NIA-funded Alzheimer’s Disease Research Centers (ADRCs) recruiting and collecting data on subjects with cognitive function ranging from normal to dementia. The NACC Uniform Data Set (UDS) is a large longitudinal dataset that includes demographic and standardized clinical data collected approximately annually. All test centers administered standardized forms and informed consent was collected from all subjects and their informants. Detailed information on the cohort and neuropsychological battery of tests included in UDS is described elsewhere^18-20^. NACC-UDS with a December 2018 data freeze date was used for this study.

### Patient Groupings

MBI status was derived from UDS using a published algorithm^21,22^ for transformation of the Neuropsychiatric Inventory Questionnaire (NPI-Q)^23^ items to MBI domains. Specifically, ten NPS domains from the NPI-Q were used to populate the five MBI domains of decreased motivation (NPI-Q apathy/indifference); emotional/ affective dysregulation (NPI-Q depression/dysphoria, anxiety, elation/euphoria); impulse dyscontrol (NPI-Q agitation/aggression, irritability/lability, aberrant motor behavior); social inappropriateness (NPI-Q disinhibition); and abnormal perception or through content (NPI-Q delusions, hallucinations). To obtain the MBI total score, these five transformed domain scores were added together. As the NPI-Q has a reference range of one month, to meet MBI persistence of symptoms criteria, individuals with MBI total score >0 at two consecutive annual visits were classified as MBI positive (MBI+) and their MBI scores were calculated as the average over the interval. Those with no NPS were classified as MBI negative (MBI-) for comparison. To determine subjective cognitive decline, the SCD-Initiative Workgroup criteria^22^ were used as a framework and operationalized in NACC as follows: 1) endorsement by participant of a decline in memory on the UDS B9 form; and 2) normal cognition.

**Figure 2** shows the step-by-step process for participant inclusion/exclusion. All NACC participants from 2005-2018 were initially considered for inclusion. The initial step was to classify based on MBI status (MBI+/-) and those with transient NPS not meeting MBI duration criteria were excluded. Study endpoint was chosen a priori to be 3 years to reflect clinical practice and design of observational cohort studies. This approach provided a concrete time frame to assess change, in order to balance the need to wait long enough to see change, but to also minimize attrition that would accrue due to age-related mortality and other diseases that confound cognitive assessments. We included participants with a follow-up visit ~3 years after the baseline visit to evaluate the change in Clinical Dementia Rating Scale (CDR® Dementia Staging Instrument)^24^ score over time and participants were excluded if they were missing the 3-year study visit. SCD status was then determined and participants with a baseline CDR>0 were excluded. Finally, those with a baseline diagnosis of a psychiatric illness were excluded.

**Figure 2:**
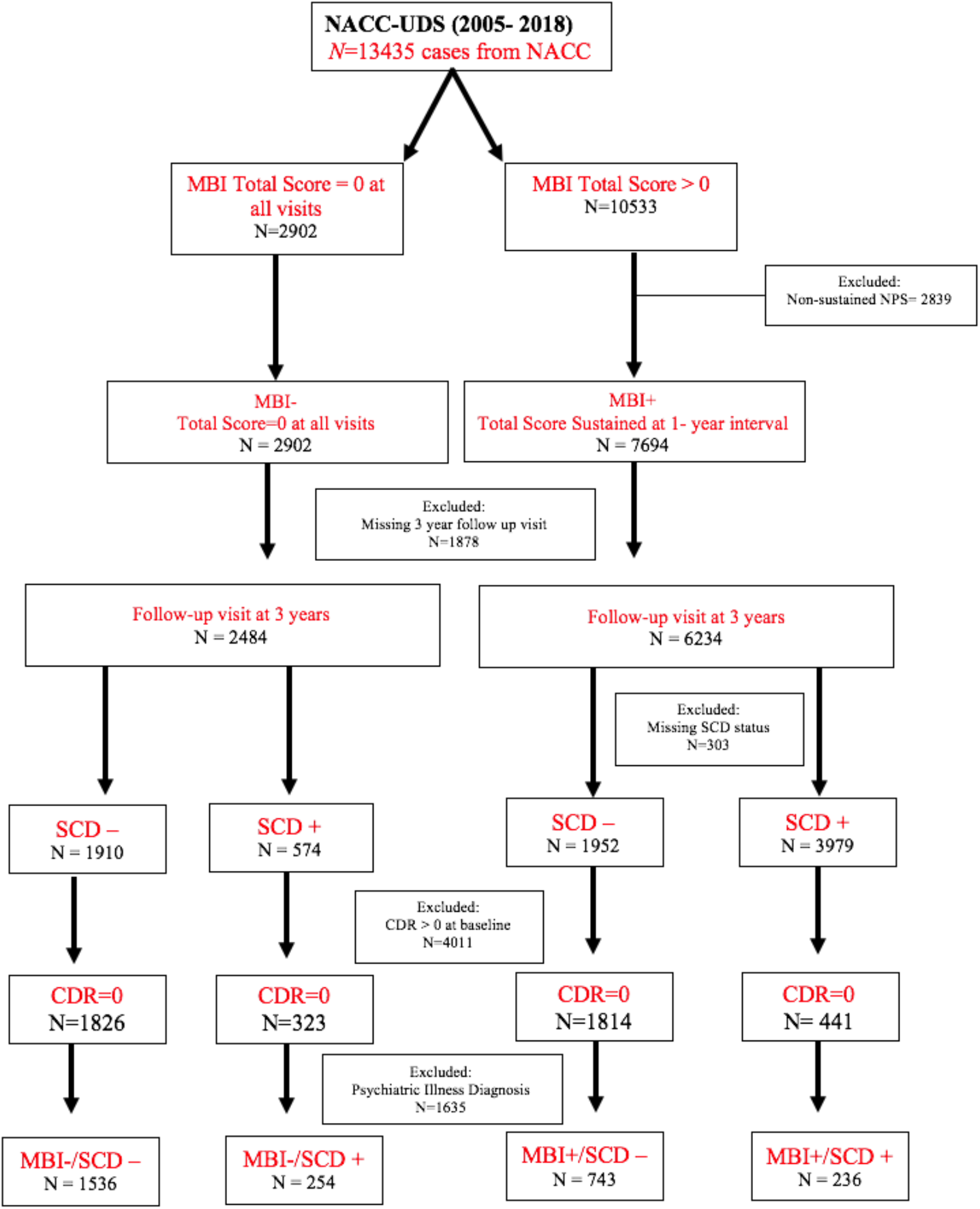
Flowchart of participants from NACC included for analysis.

### Study Variables

Baseline variables included age, sex, education, and MBI/SCD category. Our primary outcome measure, the CDR, consists of six domains: memory, orientation, judgment and problem solving, community affairs, home and hobbies, and personal care^24^. In our study, we used the global standard CDR score, which assesses the level of impairment, and ranges in severity from no impairment (CDR=0), questionable impairment (CDR=0.5 – corresponding to MCI), mild impairment (CDR=1 – corresponding to mild dementia), to moderate to severe impairment (CDR=2-3). All participants had a baseline CDR score of 0, and we measured the change in cognitive status at 3 years.

## Statistical Analysis

Categorical variables were analyzed with χ2 test, and the continuous variables were analyzed using one way ANOVA. We defined patient groups ordinally according to the absence or presence of SCD, MBI, or both at baseline. Cognitive decline was defined as CDR >0 after 3 years. We tested the ordinal by ordinal association of patient groups and cognitive decline using linear by linear association and gamma statistic, and computed odds ratios (OR) for cognitive decline using logistic regression with the patient group having neither SCD nor MBI at baseline serving as the reference group. In this model, we included terms for all variables reaching statistical significance (*p*<0.05) in the univariate analyses to calculate Adjusted Odds Ratios (AOR). All analyses were conducted in SPSS v24 (IBM Corporation) with *α* set at 0.05.

## Results

The final sample consisted of 2769 participants with CDR=0 at baseline. Participants had neither MBI nor SCD (MBI-SCD-; n=1536); SCD but no MBI (MBI-SCD+; n=254); MBI but no SCD (MBI+SCD-; n=743); and both MBI and SCD (MBI+SCD+; n=236). There were significant differences in sex, age, ethnicity, and a history of hypertension but no significant differences regarding any other clinical and demographic characteristic investigated (**Table 2**).

**Table 2.**
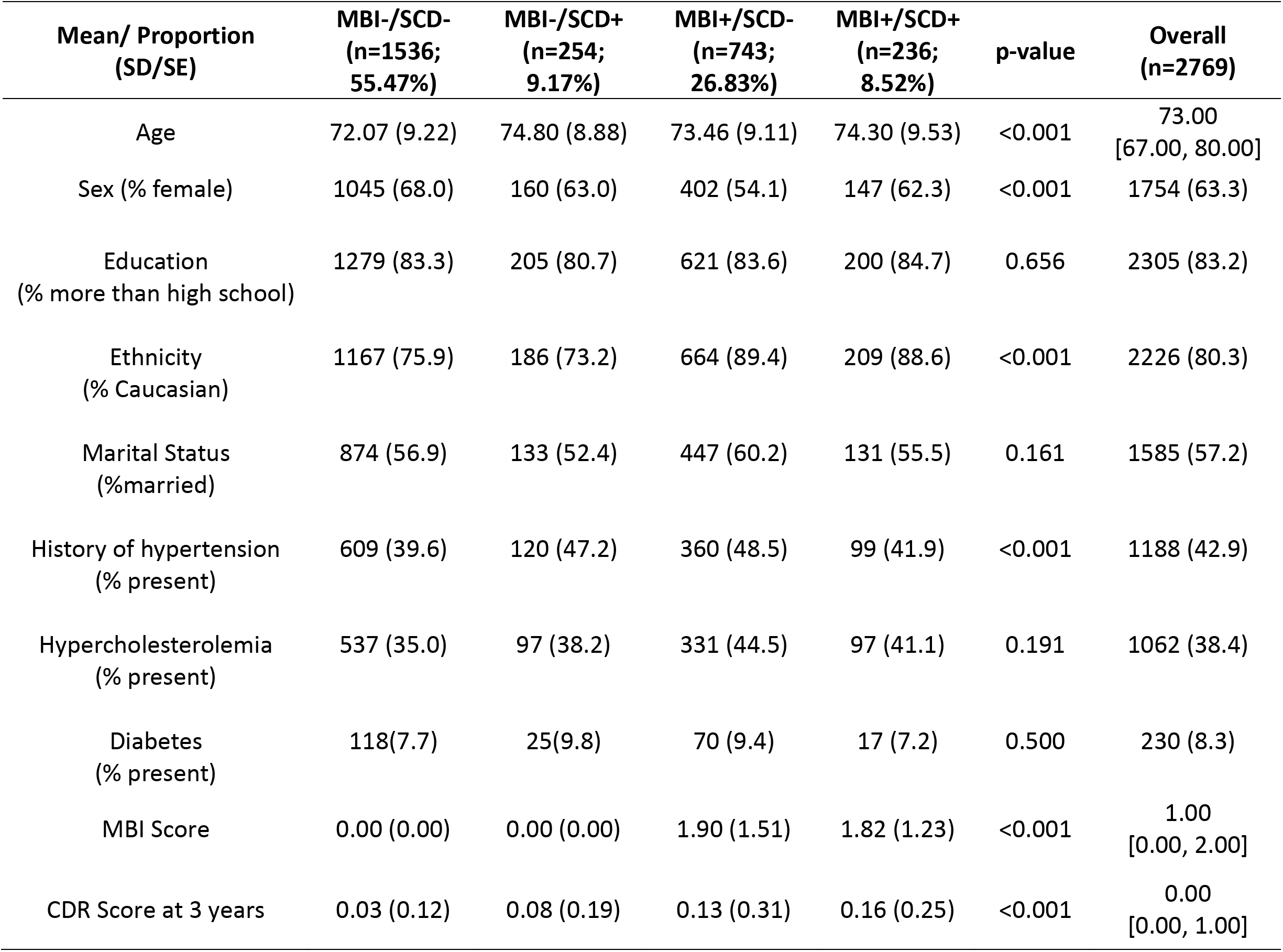
Summary statistics for demographics and MBI-C score by patient grouping in those with baseline CDR=0 (n=2769).

Over the 3 years of follow-up, 349/2769 (12.6%) individuals had evidence of cognitive decline. In **Figure 3A**, we present the incidence of cognitive decline according to the baseline presence of MBI, SCD, or their combination. Of the 1536 MBI-SCD-participants, 80 (5.21%) progressed to CDR>0 at 3 years, while progression for MBI-SCD+ was 42/254 (16.54%), MBI+SCD-was 154/743 (20.73%), and MBI+SCD+ was 73/236 (30.93%). This highly significant difference (linear-by-linear=193.24, df=1, *p*<0.001) also revealed a strong ordinal by ordinal symmetry (gamma=0.56, SE=0.031, Approximate T=12.62, *p*<0.001).

**Figure 3:**
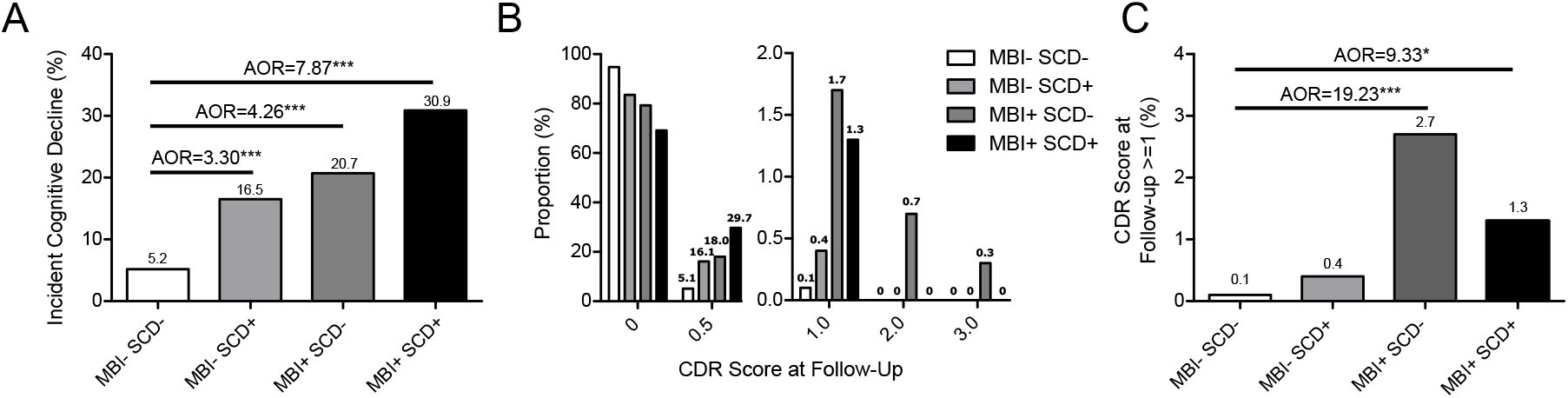
a) Odds of CDR>0 after three years vs. MBI/SCD grouping; b) CDR score at follow up (baseline CDR=0); c) Odds of dementia (CDR≥1) after 3 years vs. MBI/SCD grouping. Adjusted Odds Ratios (AORs) are adjusted for age, sex, ethnicity and hypertension.

To quantify the increased risk of incident cognitive decline according to these baseline risk definitions, we used logistic regression to generate adjusted odds ratios (AOR) and 95% confidence intervals (CI). The odds of change to CDR>0 was 8.15 times higher for MBI+SCD+ than MBI-SCD-(95%CI 2.42-5.38, *p*<0.001; AOR=7.87, 95%CI: 5.46-11.35, *p*<0.001). Those with MBI+SCD-had 4.76 times the odds of increased CDR than MBI-SCD-individuals (95%CI 3.57-6.34, *p*<0.001; AOR=4.26, 95%CI: 3.17-5.73, *p*<0.001). Those with MBI-SCD+ had 3.76 times the odds of increased CDR than MBI-SCD-individuals (95%CI 5.71-11.64, *p*<0.001; A0R=3.30, 95%CI: 2.20-4.96, *p*<0.001). Covariates for all models were age, sex, ethnicity, and history of hypertension.

We then examined the distribution of CDR scores at follow-up according to the presence of SCD, MBI, or both at baseline (**Figure 3B**), which revealed that the magnitude of progression from CDR 0 increased incrementally according to baseline characteristics. This distribution in CDR scores was significantly different across groups (linear-by-linear=165.96, df=1, *p*<0.001), with strong ordinal by ordinal symmetry (gamma=0.69, Approximate T=4.40, *p*<0.001).

Over the 3 years of follow-up, 2/1536 of the MBI-SCD-participants (0.1%) progressed to CDR≥1 (dementia), compared to 1/254 of the MBI-SCD+ participants (0.4%), 20/743 of the MBI+SCD-participants (2.7%), and 3/236 of the MBI+SCD+ participants (1.3%) (**Figure 3C**). To quantify this increased risk of incident dementia over the follow-up period, we used logistic regression. Compared to MBI-SCD-, the odds of progressing to dementia over the follow-up period were 21.21 (95%CI: 4.94-91.01, *p*<0.001; AOR=19.23, 95%CI: 4.40-84.03, *p*<0.001) for MBI+SCD- and 9.87 (95%CI: 1.64-59.41, *p*<0.05; AOR=9.33, 95%CI: 1.53-56.78, *p*<0.05) for MBI+SCD+ individuals. Covariates for all models were age, sex, ethnicity, and history of hypertension.

## Discussion

In an analysis of a longitudinal cohort of 2769 participants, we demonstrated that those who are cognitively unimpaired, have MBI, SCD, or both MBI and SCD lie on a continuum of risk for incident cognitive decline and dementia. Importantly, MBI was associated with progression to CDR >0 at the three year visit even when cognitive symptoms were absent *(i.e*. in the absence of SCD). The OR for progression to CDR >0 was numerically higher in persons with MBI alone (20.7% progression rate; OR 4.76) compared with SCD alone (16.5% progression rate; OR 3.76) but this different was not statistically significant. MBI was also associated with progression from normal cognition to dementia (CDR≥1), with very high ORs (OR of 19.23 for MBI alone and 9.87 for MBI+SCD+) but these analyses were based on only a handful of events.

In the new NIA-AA research framework for AD, stage 2 is a distinct transitional stage between stage 1 (asymptomatic) and stage 3 (mildly impaired). This framework is termed *clinical* staging rather than cognitive staging to recognize that early clinical manifestations of AD may be either cognitive or neurobehavioral^17^. As per the framework “an individual may be placed into stage 2 on the basis of neurobehavioral symptoms alone, that is, without evident cognitive decline”. Importantly, “mild neurobehavioral changes—for example, changes in mood, anxiety, or motivation—may coexist. In some individuals, the primary complaint may be neurobehavioral rather than cognitive. Neurobehavioral symptoms should have a clearly defined recent onset, which persists and cannot be explained by life events”^17^. The acknowledgement of early neurobehavioral symptoms in the NIA-AA framework is consistent with the conceptualization of MBI by the 2016 International Society to Advance Alzheimer’s Research and Treatment – Alzheimer’s Association (ISTAART-AA) MBI criteria^9^. The MBI criteria mandate that NPS emerge in later life and represent a change from longstanding patterns of behaviour (i.e. NIA-AA stage 2 “*clearly defined recent onset*”), persist for ≥6 months (i.e. NIA-AA stage 2 “*which persists*”), and cannot be better accounted for by medical or psychiatric conditions, including life stressors (i.e. NIA AA stage 2 “*cannot be explained by life events*’”).

We excluded participants with a psychiatric diagnosis at baseline as MBI diagnosis is precluded by the presence of a psychiatric disorder. In advance of cognitive changes, there is conflicting evidence regarding the links between psychiatric history and dementia risk^25,26^. The confounds in the study of the relationship between psychiatric symptoms and incident dementia have in part been due to a psychiatric nosology that is silent on the natural history of symptoms. However, several large longitudinal cohorts have provided compelling evidence that the age of onset of psychiatric symptoms is a crucial factor in determining the nature of these symptoms. These studies suggest that the later in life the onset of psychiatric symptomatology, the more likely these symptoms represent the early stages of a neurodegenerative process, that precede dementia by 5-11 years^27-30^. In a 5-year study of older adults, MBI had a higher conversion rate to dementia than a psychiatric comparator group consisting of late life psychiatric disorders^11^. From a community cohort of 9,931 participants, the emergence of MBI was associated with decline in attention and working memory at 1 year^10^. In psychiatry and neurology specialty clinic samples, incidence of dementia was higher for MBI than other psychiatric disorders^11,12^. This evidence suggests that chronic and recurrent psychiatric symptoms reflect a psychiatric disorder framed in the context of psychiatric conditions, sometimes *neurodevelopmental* in origin, supporting exclusion from our analysis. In contrast late onset psychiatric symptoms may be prodromal or precursor to cognitive decline and dementia, framed in the context of *neurodegeneration*. The ISTAART-AA MBI criteria were developed with an appreciation of the difference between pre-dementia behavioral changes and psychiatric disease in later life^9^, and are now harmonized with the biological understanding of AD in the NIA-AA clinical staging framework.

Neurobiological evidence supports the association between MBI and neurodegenerative disease. For example, cortical amyloid has been shown to moderate the association between worsening depressive symptoms and declining cognition^31^, while subcortical amyloidosis has been associated with anxiety in cognitive normals^32^. Studies have linked MBI with known AD genetic loci^13^, and have demonstrated that the relationship between AD genetic risk and cognition in healthy older adults is stronger in individuals with MBI^14^. Using machine learning models, MBI was determined to be as powerful as hippocampal volume in predicting dementia diagnosis 40 months later in a group of non-demented older adults^33^. In a study of 96 older adults with normal cognition, greater MBI burden was associated with higher PET amyloid signal, which was not dependent on the presence or absence of SCD^16^. Longitudinal data has also linked the validated biomarker neurofilament light (NfL) with MBI. In a study of 584 non-demented patients, MBI was associated with faster plasma NfL accumulation over 2 years compared to patients without MBI, with time*MBI status the only significant interaction to predict change in NfL concentration^15^. These biological links between MBI and known dementia mechanisms, further link MBI and NIA-AA clinical stage 2 disease. Thus, an increasing body of evidence suggests that MBI is likely a consequence of emerging dementia proteinopathies, manifesting independently of cognitive symptoms.

SCD is also represented in Stage 2 AD of the NIA-AA research framework in which there is subjective or objective evidence of subtle decline, not meeting criteria for objective impairment. On the AD continuum, subjective complaints of cognitive impairment would be considered evidence of subtle cognitive decline as transitional to a more impaired stage, and attributable to the pathologic process^17^. Meta-analysis of large longitudinal cohorts has shown that SCD is associated with ORs of 6 of progression to MCI and 2 for progression to dementia over a mean of 4.8 years^6^. In one study, up to 74% of people aged 70 years and older who performed normally on standard cognitive tests self-reported a subjective decline in cognitive functioning. Of these 74%, 14% represented a group at higher risk of incident MCI^34^. In a study of older adults with SCD, ascertained using a composite score of 3 rating scales, 26% were determined to be Aβ+^5^. However, significant inter-site variability in the association between SCD and abnormal cerebrospinal fluid amyloid levels has been attributed to different recruitment approaches and a lack of standardized case definitions and ascertainment^35^. There can be other contributors to subjective complaints of cognitive decline. Medical issues, stressors and even medications (such as those with an anticholinergic burden) can result in subtle cognitive impairment in the absence of neurodegeneration, which might compromise the specificity of the SCD construct. Nonetheless, as with MBI, SCD reflects emerging dementia proteinopathies associated with AD and is the index clinical manifestation of the neurodegenerative process for some.

The source of information for MBI status in our study was the NPI-Q^23^ completed by an informant. The NPI-Q was developed to measure NPS in dementia, and the symptoms as described are relevant to an aging population with neurodegenerative disease. Informant reports have shown to be more reliable assessments of NPS in neurodegenerative disease to minimize the impact of anosognosia^36^. Coincidentally, in another study of SCD, confirmation of decline by an informant was the best predictor of worse cognitive performance and lower gray matter volumes^37^. Anosognosia is also important to consider in the assessment of SCD. The INSIGHT-PreAD study showed that patients with low cognitive awareness (subjects reporting fewer difficulties than their relatives do) showed greater amyloid burden and lower cortical metabolism, compared to the high awareness group^38^. Thus, self-report of symptoms alone, whether cognitive or behavioral, may not be adequate to capture early disease.

MBI and SCD intersect in some instances and may measure similar features in different ways. For example, a study of SCD determined that *worries* about self-perceived functioning were associated with Ab positivity, rather than subjective cognitive functioning itself^39^. Worries or concerns are included in the SCD plus criteria, proposed to increase specificity for detecting preclinical AD^3^. Worry is also a component of the MBI *affective dysregulation* domain, which includes emergent mood and anxiety symptoms. A large Korean cohort study found a similar result with SCD and depressive symptoms (also a component of MBI *affective dysregulation*): SCD and depressive symptoms were both independent predictors of dementia, but together contributed to dementia development through their interaction^40^. The approach to psychiatric symptomatology in SCD has generally utilized traditional constructs of personality (e.g. neuroticism) and psychiatric conditions^41-43^. However, a change in personality to greater neuroticism (which is a *neurodevelopmental* construct) can also be framed as the emergence of MBI *affective dysregulation*, if considered in a *neurodegenerative* frame of reference^41^. This intersection of MBI and SCD is consistent with both constructs being represented in NIA-AA stage 2 AD.

Our data indicate that in cognitively normal older adults, the neurobehavioural axis of dementia risk represented by MBI, and neurocognitive axis of dementia risk represented by SCD, have complementary associations with the risk of progression to MCI and dementia. As operationalized in our study, MBI appears to be at least as strong a risk factor for progression to MCI or dementia as SCD and the two constructs have overlapping features. The combination of both MBI and SCD was associated with the highest risk.

There are practical implications of our findings. Clinically, as a complement to screening for subjective and objective cognitive symptoms in older adults, incorporating MBI into clinical assessments may provide complementary information and better risk stratification^44^. Not infrequently, dementia patients are first given a psychiatric diagnosis when presenting with a neuropsychiatric symptom, resulting in delays to treatment^45,46^.

Identifying MBI would prompt clinicians to consider neurocognitive disorders on the differential diagnosis, and flag patients who might benefit from imaging or further workup.

These findings also have clinical trial implications. Research and development costs are higher for Alzheimer’s disease (AD) than other therapy areas due to lower success rates and longer development times^2^. Despite the fact that changes in brain structure and function occur up to 20 years before gross memory impairment^47^, screening for preclinical disease is expensive and inefficient, usually requiring detailed neuropsychological testing, and amyloid/tau PET imaging or CSF analysis (or both). Leveraging the ease of measurement of MBI, in conjunction with SCD, could be an inexpensive and scalable method to select patients at highest risk for biomarker positivity and cognitive decline. For dementia prevention trials, combining MBI and SCD can increase yield and improve signal-to-noise ratios for clinical trial screening. This reduction in screen failures can increase trial efficiency and decrease trial cost.

## Limitations

The NACC participants are mostly white, highly educated, volunteers seeking care and consultation at urban, university-based centers, and the finding may not generalize to other settings. For MBI case ascertainment, we used the NPI-Q^23^ rather than the validated MBI checklist (MBI-C)^48-50^ which was developed specifically to diagnose MBI. However, we used a validated algorithm to convert NPI-Q scores to MBI-C scores and required NPS at 2 consecutive visits to match the MBI criterion of new, persistent symptoms.

## Conclusions

We have demonstrated that MBI, a neurobehavioural syndrome, is an important predictor of incident cognitive decline at 3 years in cognitively normal subjects supporting the use of MBI as a powerful risk assessment tool. Our findings suggest that MBI is at least as useful as SCD in assessing risk for incident cognitive decline and dementia, and that the two constructs are likely complementary. Assessment of the neurobehavioral and neurocognitive axes at the same time are required in cognitively normal individuals to better define their risk. Our findings add support to the inclusion of later life emergent and persistent neurobehavioural symptoms in the NIA-AA research framework for AD.

## Data Availability

Data is available upon request.

## Acknowledgements

Grant support included the Alzheimer Society of Calgary via the Hotchkiss Brain Institute (Ismail) and the Canada Research Chairs program (Forkert). We also acknowledge the Mathison Centre for Mental Health Research & Education, and the Ron and Rene Ward Centre for Healthy Brain Aging for support.

Data used in this study was from the NACC database which is funded by NIA/NIH Grant U01 AG016976. NACC data are contributed by the NIA-funded ADCs: P30 AG019610 (PI Eric Reiman, MD), P30 AG013846 (PI Neil Kowall, MD), P30 AG062428-01 (PI James Leverenz, MD) P50 AG008702 (PI Scott Small, MD), P50 AG025688 (PI Allan Levey, MD, PhD), P50 AG047266 (PI Todd Golde, MD, PhD), P30 AG010133 (PI Andrew Saykin, PsyD), P50 AG005146 (PI Marilyn Albert, PhD), P30 AG062421-01 (PI Bradley Hyman, MD, PhD), P30 AG062422-01 (PI Ronald Petersen, MD, PhD), P50 AG005138 (PI Mary Sano, PhD), P30 AG008051 (PI Thomas Wisniewski, MD), P30 AG013854 (PI Robert Vassar, PhD), P30 AG008017 (PI Jeffrey Kaye, MD), P30 AG010161 (PI David Bennett, MD), P50 AG047366 (PI Victor Henderson, MD, MS), P30 AG010129 (PI Charles DeCarli, MD), P50 AG016573 (PI Frank LaFerla, PhD), P30 AG062429-01(PI James Brewer, MD, PhD), P50 AG023501 (PI Bruce Miller, MD), P30 AG035982 (PI Russell Swerdlow, MD), P30 AG028383 (PI Linda Van Eldik, PhD), P30 AG053760 (PI Henry Paulson, MD, PhD), P30 AG010124 (PI John Trojanowski, MD, PhD), P50 AG005133 (PI Oscar Lopez, MD), P50 AG005142 (PI Helena Chui, MD), P30 AG012300 (PI Roger Rosenberg, MD), P30 AG049638 (PI Suzanne Craft, PhD), P50 AG005136 (PI Thomas Grabowski, MD), P30 AG062715-01 (PI Sanjay Asthana, MD, FRCP), P50 AG005681 (PI John Morris, MD), P50 AG047270 (PI Stephen Strittmatter, MD, PhD).

